# Daily Rapid Antigen Testing in a University Setting to Inform COVID-19 Isolation Duration Policy

**DOI:** 10.1101/2022.03.11.22272264

**Authors:** Rebecca Earnest, Christine Chen, Chrispin Chaguza, Nathan D. Grubaugh, Madeline S. Wilson, the Yale COVID-19 Resulting and Isolation Team

## Abstract

**Importance:** The suitability of the currently recommended 5-day COVID-19 isolation period remains unclear in an Omicron-dominant landscape. Early data suggest high positivity via rapid antigen test beyond day 5, but evidence gaps remain regarding optimal isolation duration and the best use of limited RATs to exit isolation.

**Objective:** To determine the percentage of SARS-CoV-2 infected persons who remain positive via RAT on isolation day 5+ and assess possible factors associated with isolation duration.

**Design:** We evaluated daily rapid antigen test case series data from 324 persons in a managed isolation program who initially tested positive between January 1 and February 11, 2022, an Omicron-dominant period. Arrival tests and twice-weekly screening were mandated. Positive persons isolated and began mandatory daily self-testing on day 5 until testing negative. Trained staff proctored exit testing.

**Setting:** A mid-sized university in the United States.

**Participants:** University students in isolation.

**Main Outcomes and Measures:** The percentage of persons remaining positive on isolation day 5 and each subsequent day. The association between possible prognostic factors and isolation duration as measured by event-time-ratios (ETR).

**Results:** We found 47% twice-weekly screeners and 26-28% less frequent screeners remained positive on day 5, with the percentage approximately halving each additional day. Having a negative test ≥ 10 days before diagnosis (ETR 0.85 (95% CI 0.75-0.96)) and prior infection > 90 days (ETR 0.50 (95% CI 0.33-0.76)) were significantly associated with shorter isolation. Symptoms before or at diagnosis (ETR 1.13 (95% CI 1.02-1.25)) and receipt of 3 vaccine doses (ETR 1.20 (95% CI 1.04-1.39)) were significantly associated with prolonged isolation. However, these factors were associated with duration of isolation, not infection, and could reflect how early infections were detected.

**Conclusions and Relevance:** A high percentage of university students during an Omicron-dominant period remained positive after the currently recommended 5-day isolation, highlighting possible onward transmission risk. Persons diagnosed early in their infections or using symptom onset as their isolation start may particularly require longer isolations. Significant factors associated with isolation duration should be further explored to determine relationships with infection duration.

**Key Points:** *Question:* What percentage of SARS-CoV-2 infected persons remain positive via rapid antigen test on days 5+ of isolation?

*Findings:* In this case series of 324 university students, 47% of twice-weekly screeners and 26-28% of less frequent screeners remained positive via rapid antigen on isolation day 5, with the percent still positive approximately halving with each subsequent day.

*Meaning:* While isolation duration decisions are complex, our study adds to growing evidence that a 5-day isolation may be 1-2 days too short to sufficiently reduce the onward transmission risk, particularly for those in dense settings or among vulnerable populations.

## Introduction

In December 2021, the Centers for Disease Control and Prevention (CDC) reduced the recommended COVID-19 isolation period for the general population from 10 to 5 days following the symptom onset or a positive viral test.^1^ To end isolation, persons must have resolving symptoms and wear a mask for an additional 5 days; however, a negative exit test was not required. The rationale for the shortened isolation is based on practical and scientific considerations, namely weighing the societal and economic burdens against the diminishing risk of onward transmission as a positive person proceeds through their infection. The CDC revised its guidelines as Omicron rapidly grew to dominance in the United States, increasing from 1% to > 50% of reported sequences over a 2-week period in December 2021.^2^ Early analysis suggests potentially different viral dynamics for Omicron versus Delta, with lower peak viral RNA and shorter clearance periods for Omicron versus Delta, but similar proliferation times and clearance rates.^3^ As the current recommendations based on estimates for earlier SARS-CoV-2 variants, more data are needed to understand their appropriateness for Omicron.

Additionally, the updated guidance acknowledges the possibility of onward transmission following a 5-day isolation, citing an earlier United Kingdom modeling study estimating that 31% of persons remain infectious on day 5.^4^ Emerging literature on exit testing from an Omicron-dominant period further indicates that high proportions of persons remain potentially infectious beyond day 5. Studies of managed isolation programs through schools or employers found positivity via rapid antigen test (RAT) on days 5-9 ranging from 31-58%, although daily testing among all persons was not conducted.^5–7^ A near-daily PCR testing study reported a day 5 positivity of ranging from 39-52%.^3^ However, key gaps in our understanding persist. Firstly, some studies did not test all persons daily from day 5 and are right-censored (i.e. some persons did not test negative). In addition, non-mandatory testing raised the possibility of selection bias, with sicker persons potentially opting not to exit test. Lastly, most studies did not adjust for the approximate time of infection.

While PCR tests are a preferred initial diagnostic option due to their high sensitivity, RATs are more suitable for exit testing when the goal is to determine when a person is likely no longer infectious. High PCR sensitivity may result in positive tests beyond the infectious period, leading to unnecessarily long isolations.^8,9^ RAT positivity is generally associated with culturable virus, which itself is often a proxy for infectiousness.^9–11^ To investigate concerns that RATs may have inferior performance for Omicron versus Delta infections, a study compared same-day positivity between the variants, finding similar sensitivity of RAT and PCR tests.^12^ Lastly, RATs have the advantage of relative affordability, fast turnaround time, and at-home self-administration compared to PCR, making them the only viable exit test option for much of the population.^13,14^

To address the evidence gaps regarding optimal isolation durations and how SARS-CoV-2-positive persons can best utilize exit RATs when resources are limited, we evaluated daily RAT data from 324 persons in a university-managed isolation program who initially tested positive for SARS-CoV-2 between January 1 and February 11, 2022. We posed two questions: 1) what percentage of SARS-CoV-2-positive persons remained positive via RAT on day 5 of isolation and each subsequent day until testing negative and 2) what factors were associated with isolation duration?

## Methods

The university required undergraduate students to screen upon campus arrival and subsequently twice-weekly on designated days. SARS-CoV-2 positive students isolated and participated in mandatory daily rapid antigen self-testing beginning on day 5 following diagnosis (the CDC’s recommended end of isolation^1^) until they tested negative. Diagnosis (day 0) was defined as the earliest positive or inconclusive test date. All inconclusive persons subsequently tested positive. Trained staff observed the exit testing process and confirmed the result. Upon testing negative, students ended isolation but continued mandatory masking until day 10. All testing was conducted using the Quidel QuickVue At-Home OTC COVID-19 Test, a lateral flow immunoassay that qualitatively detects the SARS-CoV-2 nucleocapsid protein antigen.^15^ The test received a FDA Emergency Use Authorization for prescribed home use with self-collected anterior nares swab specimens and has a sensitivity of 84.8% (95% CI 71.8-92.4) and a specificity of 99.1% (95% CI 95.2-99.8).

We used RStudio v1.4.1106 for our analyses.^16^ We calculated the percent still positive as the number of positive persons each day divided by the total number of positive persons. To assess prognostic factors associated with the time to event (i.e. testing negative), we coded an accelerated failure time (AFT) lognormal regression model using the *survival* R package v3.2-13.^17,18^ We selected the AFT model due to its suitability for interval-censored data.^19^ Since students enter the study on day 0 but are not rapid tested until day 5, any persons testing negative on day 5 are interval-censored, with their true negative time falling between day 1 and day 5. We compared model fits using various distributions and selected the fit resulting in the lowest Akaike Information Criterion value. We exponentiated the regression coefficients to calculate the event-time-ratio (ETR), which are associated with prolonged isolation duration when > 1 and decreased duration when < 1. We checked the assumption that the ratio of survival times (i.e. the event-time-ratio) is constant for all fixed probabilities of S(t), the survival function, by a visual inspection of QQ plots generated for each covariate level comparison using the R package *AFTtools* v0.2.1.^20^

## Results

Our study population comprised primarily 18-22 year old students living in university dormitory housing (N=323, **Table 1**). Among them, 63% self-reported symptoms prior to or at diagnosis. We did not track symptoms beyond diagnosis, although 18/205 symptomatic persons had a symptom onset date 1 day post-diagnosis, potentially reflecting when they received their results and discussed symptoms. We found that 7% had a confirmed SARS-CoV-2 infection > 90 days before their recent diagnosis. The university did not screen asymptomatic persons with an infection ≤ 90 days due to the false positive risk. We categorized vaccinations into 1-4 doses. Generally, a non-mRNA vaccine primary series counted as 1 dose, a mRNA vaccine primary series as 2 doses, and a booster as an additional dose. Two individuals reported receiving 2 boosters, resulting in 4 doses. Only doses administered ≤ 14 days prior to diagnosis were counted toward the total.^21^ The breakdown of doses was as follows: 3% of persons had 1 dose, 27% 2 doses, 68% 3 doses, 1% 4 doses, and 2% of persons had missing data. **eTable 1** presents detailed vaccination history. Isolation duration is dependent on where a person is in their infection course when they are diagnosed. To address this, we used the time since the last negative test as an approximation of the time since infection, with 56% of persons testing negative ≤ 4 days prior to diagnosis, 15% 5-9 days prior, and 29% ≥ 10 days prior. 1 person had missing data. The ≤ 4 days group represents students compliant with university twice-weekly screening policy, the 5-9 day group a mix of non-compliant routine screeners and arrival screeners, and the ≥ 10 day group arrival screeners.

**Table 1:** Characteristics of study population completing isolation stratified by days since last negative test category. N=323. Number of persons and column percentages for each characteristic. Category totals may not add to 100% due to rounding errors.

| Characteristic |  | Number (%) |  |  |  |  |
| --- | --- | --- | --- | --- | --- | --- |
|  |  | Days Since Last Negative Test |  |  |  | Total |
| | | $\leq 4$<br>(N=181) | 5-9<br>(N=48) | $\geq 10$<br>(N=93) | Unknown<br>(N=1) | -<br>(N=323) |
| Self-Reported<br>Symptoms Prior to<br>or At Diagnosis | No | 51 (28) | 17 (35) | 46 (49) | 1 (100) | 115 (36) |
|  | Yes | 130 (72) | 29 (60) | 46 (49) | 0 (0) | 205 (63) |
|  | Unknown | 0 (0) | 2 (4) | 1 (1) | 0 (0) | 3 (1) |
| Prior Infection > 90<br>days | No | 171 (94) | 46 (96) | 84 (90) | 1 (100) | 302 (93) |
|  | Yes | 10 (6) | 2 (4) | 9 (10) | 0 (0) | 21 (7) |
| Number of Vaccine<br>Doses | 1 | 3 (2) | 0 (0) | 6 (6) | 0 (0) | 9 (3) |
|  | 2 | 38 (21) | 16 (33) | 31 (33) | 1 (100) | 86 (27) |
|  | 3 | 136 (75) | 30 (62) | 54 (58) | 0 (0) | 220 (68) |
|  | 4 | 1 (1) | 1 (2) | 0 (0) | 0 (0) | 2 (1) |
|  | Unknown | 3 (2) | 1 (2) | 2 (2) | 0 (0) | 6 (2) |

To calculate the percent still positive on day 5+, we dropped 1 person with an unknown last negative test time and 7 persons who initially tested inconclusive but used the subsequent positive test date as the isolation start, resulting in a final dataset of 315 persons. Among twice-weekly screeners, 47% of all diagnosed (N=177) remained positive on day 5, 22% on day 6, 8% on day 7, and 1-2% on days 8-13 (**Figure 1A**). Among students last testing negative 5-9 days before diagnosis, 28% of all diagnosed (N=47) remained positive on day 5, 17% on day 6, 6% on day 7, and 2-4% on days 8-9 (**Figure 1B**). Students last testing negative ≥10 days prior (N=91) experienced similar daily positivity (**Figure 1C**). A density plot of these data shows isolation duration distributions for all groups centered on day 5, with relatively larger peaks for the ≤ 4 day category on days 6+ (**Figure 1D**).

**Figure 1:**
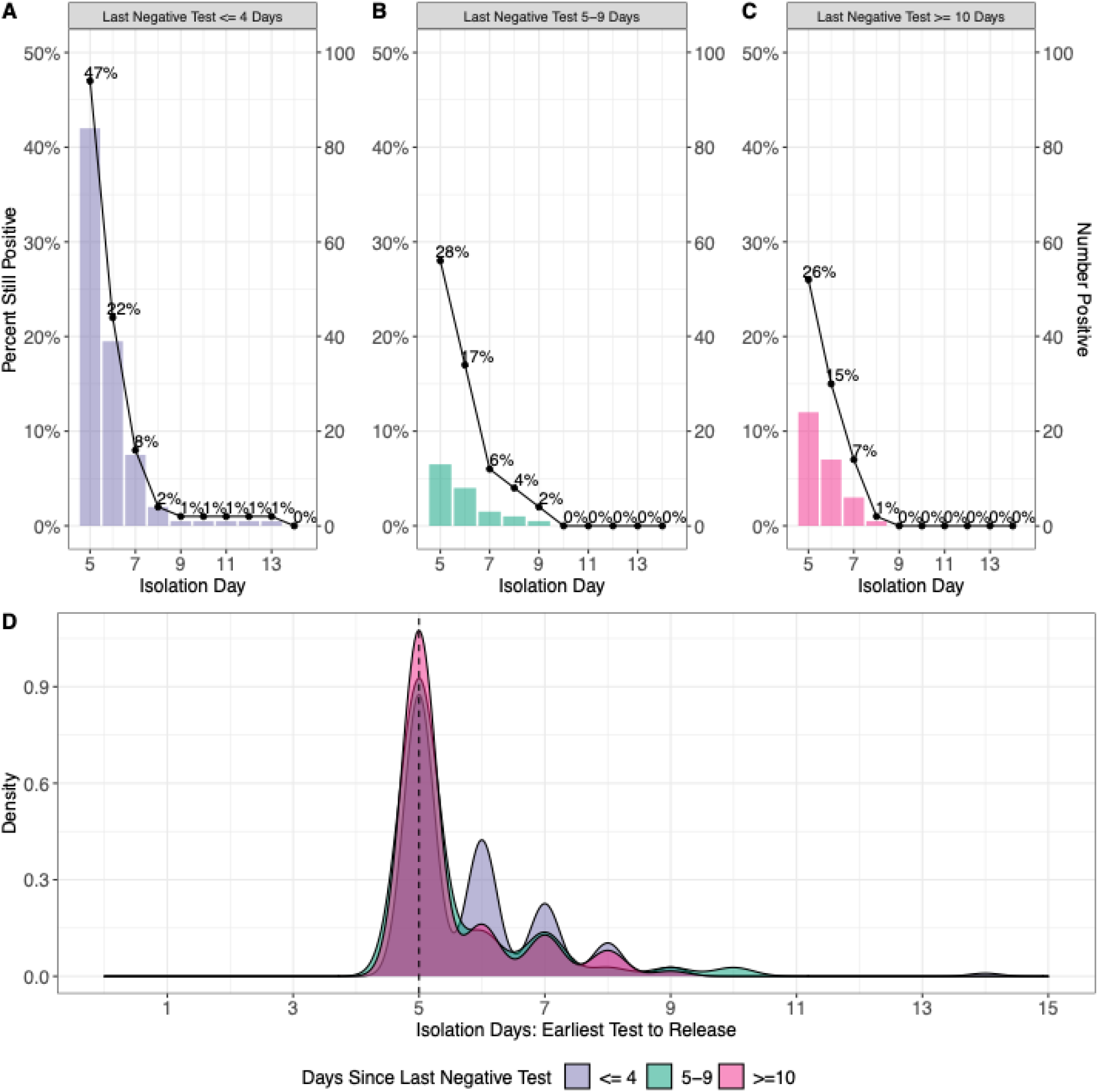
Rapid antigen testing results by isolation day and isolation duration by days since the last negative test category. Percent still positive of the original study population on the left axis and the number of positive persons on each isolation day on the right axis. 1 person was removed due to missing last negative test data and 10 persons were removed due to testing inconclusive initially but counted the first positive test as day 0. Last negative test **A)** ≤ 4 days (N=177), **B)** 5-9 days (N=47), and **C)** ≥ 10 days prior to the earliest test (inconclusive or positive) (N=91). **D)** Isolation duration (time from earliest test to test negative) by time since last negative test category. The vertical line indicates day 5, the first day of rapid antigen testing.

To evaluate possible prognostic variables for isolation duration, we conducted a survival analysis using an AFT lognormal regression model. We subset the dataset used in **Figure 1** to exclude those with 1 (N=8), 4 (N=2), or an unknown (N=6) number of vaccine doses due to small category sizes, a missing PCR cycle threshold (CT) value at diagnosis due to an external test or RAT (N=27), a missing symptom status (N=2), and those who received an international vaccine (N=8), resulting in a final sample of N=263 persons. We included time since the last negative test category as a covariate to account for possible confounding as persons in different infection stages would necessarily experience differing isolation durations. We also included symptom status, PCR CT value, and prior infection > 90 days as covariates. We created a new variable combining the number of vaccine doses (2 or 3) and the time since the last dose (< 5 months or ≥ 5 months).^22^ All “3 vaccine dose” persons except 1 received their last dose < 5 months. Finally, we included the primary series vaccine brand grouped into mRNA vaccines (Pfizer and Moderna) and J&J. **Table 2** presents the regression results and **eFigure 1** displays the isolation duration distribution for each covariate category excluding the time since the last negative test, which is displayed in **Figure 1**. We found that having a last negative test ≥ 10 days prior significantly decreased isolation duration by 15% (ETR 0.85 (95% CI 0.75-0.96)) compared to having a last negative test ≤ 4 days prior. Being symptomatic significantly increased isolation duration by 13% (ETR 1.13 (95% CI 1.02-1.25)). Having a prior infection > 90 days significantly decreased isolation duration by 50% (ETR 0.50 (95% CI 0.33-0.76)). Receipt of 3 vaccine doses significantly increased isolation duration by 20% (ETR 1.20 (95% CI 1.04-1.39)) compared to the 2 doses ≥ 5 months group. The other covariates were not significant.

**Table 2:** Event time ratios (ETR) of the association between days since last negative test, symptoms at diagnosis, CT value at diagnosis, prior infection >90 days before diagnosis, number of vaccine doses and time since last dose, and primary vaccine brand with isolation duration. N=263 persons who were fully vaccinated with Pfizer, Moderna, or J&J, did not additionally receive an international vaccine, and did not have a missing CT value or symptom status. An ETR > 1 is associated with prolonged isolation duration compared to the reference group. An ETR < 1 is associated with a decreased isolation duration.

| Covariate | Category |  | Sample Size | ETR | 95% CI | P-Value |
| --- | --- | --- | --- | --- | --- | --- |
| *Time Since Last Negative Test* | ≤4 days | ref | 155 | - | - | - |
|  | 5-9 days | - | 40 | 0.88 | (0.77-1.01) | 0.065 |
|  | ≥ 10 days | - | 68 | 0.85 | (0.75-0.96) | 0.008 |
| *Symptoms at Dx* | No | ref | 104 | - | - | - |
|  | Yes | - | 159 | 1.13 | (1.02-1.25) | 0.016 |
| *CT Value at Dx* | - | - | 263 | 1 | (0.99-1) | 0.378 |
| *Prior Infection >90 days* | No | ref | 244 | - | - | - |
|  | Yes | - | 19 | 0.5 | (0.33-0.76) | 0.001 |
| *No. Dose/Time Since Last* | 2 Doses / ≥ 5m | ref | 44 | - | - | - |
|  | 2 Doses / < 5m | - | 31 | 1.29 | (0.97-1.73) | 0.083 |
|  | 3 Doses | - | 188 | 1.2 | (1.04-1.39) | 0.012 |
| *Primary Vaccine Brand* | JJ | ref | 24 | - | - | - |
|  | mRNA | - | 239 | 1.21 | (0.89-1.65) | 0.219 |

## Discussion

We analyzed data from a mandatory, daily RAT program among university students in isolation to assess the percent still positive on day 5+ and determine possible prognostic factors for isolation duration. In addition, we approximately accounted for time since infection by stratifying our analysis by the time since the last negative test. We found a day 5 positivity of 47% in the twice-weekly screening group and 26-28% in the other less frequently screened groups (**Figures 1A-C**). For all groups, positivity approximately halved with each additional day. These results align with the expectation that more frequent screeners are diagnosed earlier in their infection, thus experiencing a longer isolation. Our findings are similar to results reported in other analyses, although most did not conduct daily mandatory testing. Studies of managed isolation programs reported RAT positivity ranging from 31-58% on days 5-9 of isolation and PCR positivity ranging from 39-52% on day 5, 25-33% on day 6, and 13-22% on day 7.^3,5–7^

Our results indicate that a 7-day isolation period is required to achieve at least a 90% probability of being negative without exit testing (**Figure 1A-C**). 5-day and 6-day isolation periods are associated, respectively, with a 53-74% and 78-85% probability of testing negative (range across last negative test categories). In cases where only a single RAT per person is available, the ideal exit test timing should be informed by the risk of onward transmission. For a person living or working in a higher-risk environment due to density, vulnerable populations, or both (e.g., university dormitories or long-term care facilities), onward transmission is potentially riskier and costlier and a day 7 exit test may be most appropriate. Persons in lower-risk environments could consider using a single RAT on day 5 and, if positive, remain in isolation until day 7. Persons with multiple RATs could begin daily exit testing on day 5. While we defined the isolation start as the initial test, CDC guidelines define it as the initial test or the symptom onset. 63% of our study population reported symptoms before or at diagnosis (**Table 1**). Symptomatic persons reported symptom onset a mean of 0.88 days before their initial test (IQR 0-1.25 days) in the last negative test ≤ 4 days group, 0.93 (IQR 0-1.25) in the 5-9 days group, and 3.04 days (IQR 0-4 days) in ≥ 10 days group. Persons using symptom onset as their isolation start accordingly may have longer isolation durations than those we measured in our study.

In our survival analysis, having a negative test ≥ 10 days, symptom status, prior infection > 90 days, and receipt of 3 vaccine doses were significantly associated with isolation duration (**Table 2**). Other covariates were not significant. For the last negative test covariate, we observed an association with shorter duration time for the ≥ 10 days and 5-9 days groups compared to the ≤ 4 days group, although only the ETR for the ≥ 10 days group was significant. The relationship between less frequent screening and shorter isolation duration is intuitive as these persons are more likely to be diagnosed later in their infection. Reporting symptoms significantly increased isolation duration. Symptomatic persons may be diagnosed earlier in their infection, even in the case of routine screening, resulting in a longer isolation period. Experiencing a prior infection > 90 days significantly decreased isolation duration. In a highly vaccinated population, having a previous infection may confer greater immunity relative to those without ^23^, reducing the isolation duration. Receipt of 3 vaccine doses was significantly associated with a longer isolation duration compared to receipt of 2 doses ≥ 5 months prior to diagnosis, an unexpected direction of effect. This finding was consistent under various formulations of the model during our exploratory phase. Another study found that vaccine boosted persons were twice as likely to test positive on an initial RAT on days 5-10 compared to unboosted persons, although not all persons tested daily.^5^ The authors suggested that boosted persons may develop symptoms earlier, leading to speedier detection and, resultantly, longer isolation durations. Accounting for the time since the last negative test in our model would likely reduce some of the bias toward earlier detection of symptomatic individuals, however this explanation remains possible. In addition, more persons in the 2 dose groups may have been infected with Delta compared to those in the 3 dose group. We observed a higher proportion of persons belonging to the 2 dose groups earlier in our study, when Delta still circulated at low levels (**eFigure 2**). If the incubation period or infection duration differ between Delta and Omicron infections, this could contribute to our findings. However, Omicron reached 97% frequency among sequenced samples in New Haven County by January 1, 2022 (with the remaining 3% composed of Delta).^24^ We observed a substantially larger sample size for the 3 dose group (N=188) compared to the 2 dose ≥ 5 (N=44) and < 5 (N=31) month groups. The larger sample may have captured more isolation duration outliers. Finally, our analysis assesses the relationship between these factors and duration of isolation, not infection. There may be other unaccounted factors associated with both the 3 dose group and isolation duration.

### Limitations

Symptom status only captures self-reported symptoms before or at diagnosis and may not always be related to the subsequent SARS-CoV-2 diagnosis. Three persons reported a symptom onset > 10 days before diagnosis. Some asymptomatic persons may have later become symptomatic. Prior infections > 90 days only included confirmed infections reported in the medical records. There likely are missed prior infections that occurred during breaks or before routine screening was implemented at the university in Fall 2021. The PCR CT value was only measured at diagnosis. Some CT values were missing due to external tests or RAT. Our study population, primarily 18-22 year old students, may not be representative of the general population due to their youth and likely lower rate of comorbidities. However, it is unlikely that older age groups or those with higher comorbidity rates would experience shorter isolation durations. We do not have a full medical history for our study population, and it is possible that some persons may experience longer isolations due to medical conditions. There could be changes in staff accuracy over time in reading RAT results, which are qualitative in nature, although their training procedures render this less likely. We do not have RAT data for days 1-4 and accounted for this interval-censoring in our analysis. RATs have a lower sensitivity than PCR, reducing the risk that a non-infectious person would remain in isolation but increasing the risk of a false negative.^8,9^ RAT positivity, while associated with culturable virus, does not mean that a person is necessarily infectious.^9–11^

## Conclusion

A high percentage (26-47%) of university students during an Omicron-dominant period remained positive after the currently recommended isolation period. Thus, the currently recommended 5-day isolation period may be too short, especially for persons using symptom onset as their isolation start or those diagnosed early in their infections. However, it is important to note that these factors are associated with duration of isolation, not infection, and thus unmeasured factors associated with diagnosis timing could affect our findings. Future research assessing daily RAT positivity beginning at the isolation start in the general population would further refine our understanding of optimal isolation periods, as would analyses characterizing what, if any, onward transmission has resulted from the current 5-day isolation period. Factors such as social distancing and masking may reduce transmission risk posed by RAT-positive persons. These considerations illustrate the complexity of recommending isolation durations for the general population, but our study adds to growing evidence that the current 5-day isolation period may be 1-2 days too short to sufficiently reduce the risk of onward transmission, especially in dense settings or ones with vulnerable populations.

## Data Availability

All original code and data have been deposited at Github and are publicly available (https://github.com/rebecca-earnest/2022_paper_isolation-rapid-antigen). Any additional information required to reanalyze the data reported in this paper is available from the corresponding author upon request.

## Acknowledgments

We thank the Yale COVID Testing and Tracing Committee and the Yale COVID-19 Resulting and Isolation Team for the substantial amount of time and effort that went into designing, implementing, and sharing data from the rapid antigen testing isolation program. This work is supported by the Yale Center for Clinical Investigation (YCCI) Multidisciplinary Pre-Doctoral Training Program. Dr. Madeline S. Wilson and Rebecca Earnest had full access to all the data in the study and take responsibility for the integrity of the data and the accuracy of the data analysis.

## Author Contributions

Conceptualization, R.E., C.Chen, N.D.G., M.S.W

Collected and provided isolation data: C.Chen, M.S.W., Yale COVID-19 Resulting and Isolation Team

Methods development, R.E., C.Chaguza

Data analysis and interpretation, R.E., C.Chaguza, N.D.G.

Supervision, N.D.G., M.S.W

Writing - Original Draft, R.E., N.D.G;

Writing - Review & Editing, all authors.

## Yale COVID-19 Resulting and Isolation Team Authors

Adam Cairns, Alyssa Cooksey, Arielle Pensiero, Asnath Mosha, Christina Pivirotto, Colleen Traub, Gabriel Velazquez, Heejun Lee, Jasper Larioza, Jelissa Neal, Jenna Bourgeois, Kathleen Donnelly, Karen Otterson, Kiley Carbone, Kristin Smith, Lauren Gillingham, Lauren Greenberg, Linda DiGangi Ehrenfels, Maen Adileh, Marie Toupou, Nathan Lubich, Nathan Yuen, Nick Davies, Nikkiyah Brown, Noor Khalid, Olivia Dumont, Onyi Umeugo, Oscar Monteagudo, Presley Hill, Quinn O’Leary, Rashea Banks, Richard Rousseau, Sam Duplantis, Sara Khalid, Saskia César, Sean Doran, Shannan Charney, Shinelle Wilkins, Sofia Martinez, Spencer Volpe, Victor Martinez Garcia, Vittoria Cappuccia

## Ethics

The Institutional Review Board from the Yale University Human Research Protection Program determined that the use of information, including information about biospecimens, is recorded by the investigator in such a manner that the identity of the human subjects cannot readily be ascertained directly or through identifiers linked to the subject and thus is exempt from IRB review of human subjects research (IRB Protocol ID: 2000032111).

## Supplementary Information

**eTable 1:**
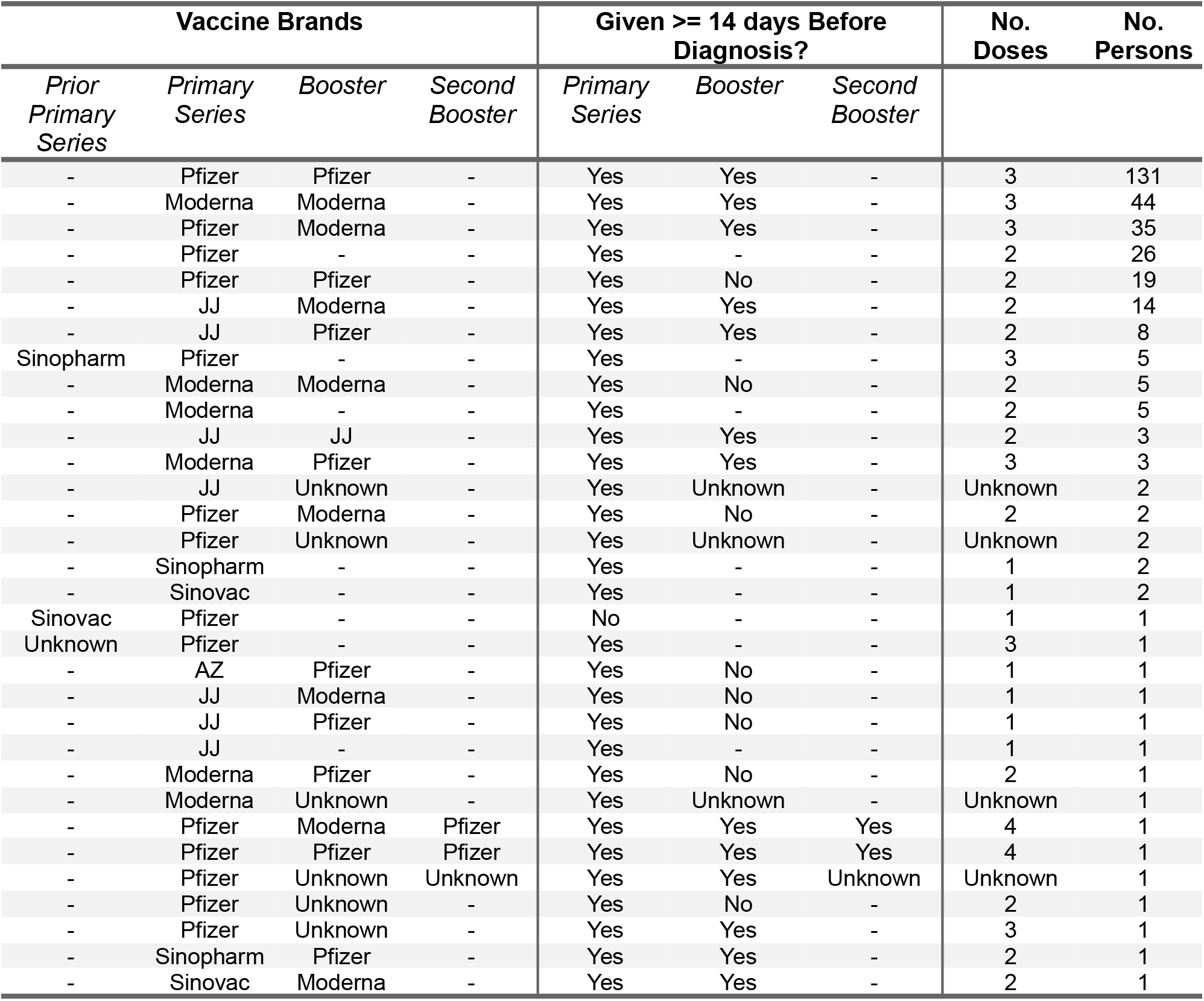
Study population vaccination history, related to Table 1. N=323. Prior primary series refers to individuals who received an international vaccine(s) prior to being fully vaccinated with a mRNA vaccine. Unknown refers to individuals with missing or unconfirmed vaccine brand or date administered data. Vaccine doses only count if fully administered at least 14 days prior to diagnosis. non-mRNA vaccine primary series correspond to one dose, mRNA vaccine primary series to two doses, and any booster as an additional dose.

**eFigure 1:**
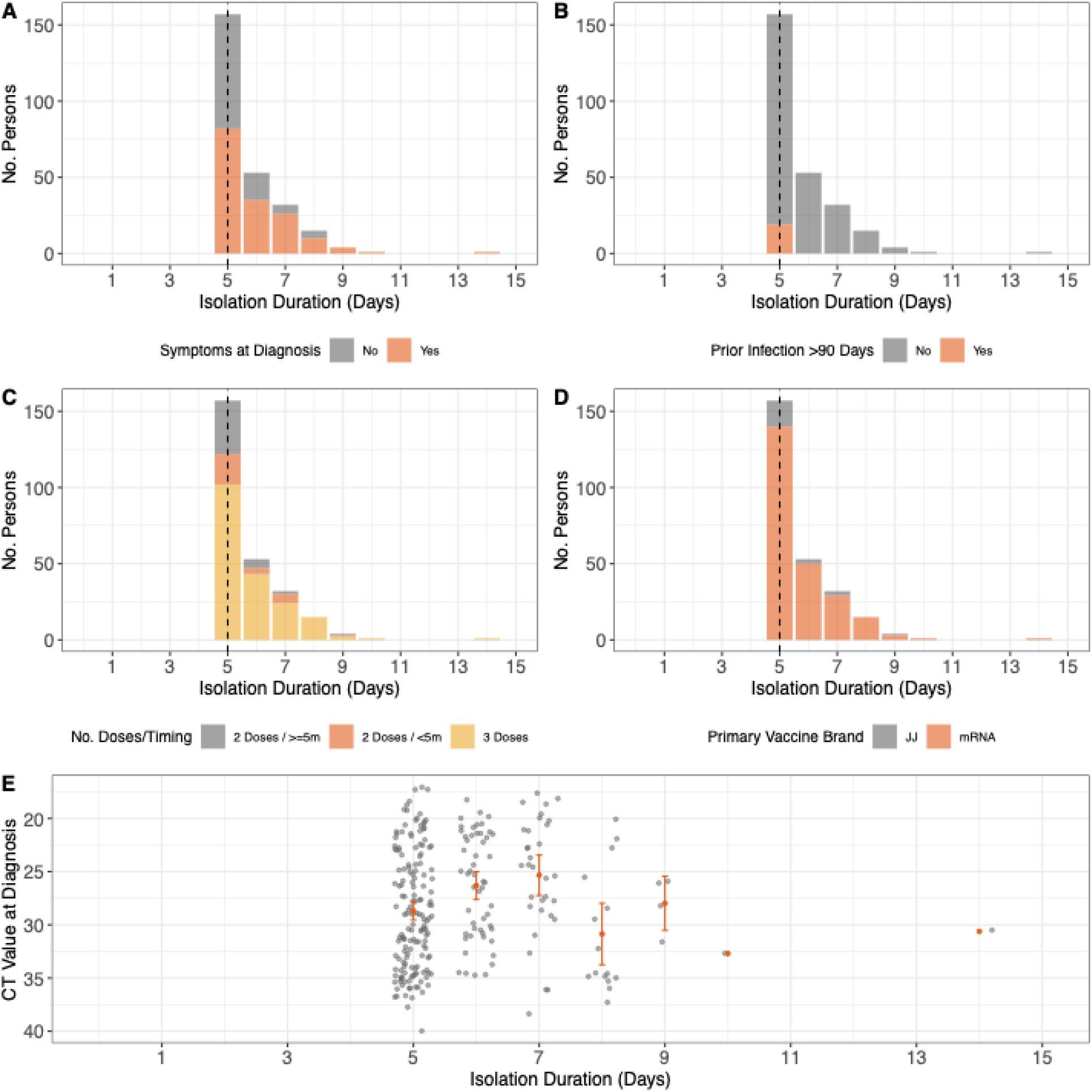
Relationship between model covariates and isolation duration, related to Table 2. N=263 persons included in the survival model analysis (Table 2). Isolation duration measured as the number of days from testing positive or inconclusive to testing negative.The vertical line indicates day 5, the first day of rapid antigen testing. We show isolation duration by A) symptom status, (B) prior infection > 90 days, C) primary vaccine brand, (D) the number of and time since the last vaccine dose, and (E) CT value at diagnosis. For (E), note the inverted y-axis as lower CT values correspond to higher viral loads. We present the mean and 95% confidence interval.

**eFigure 2:**
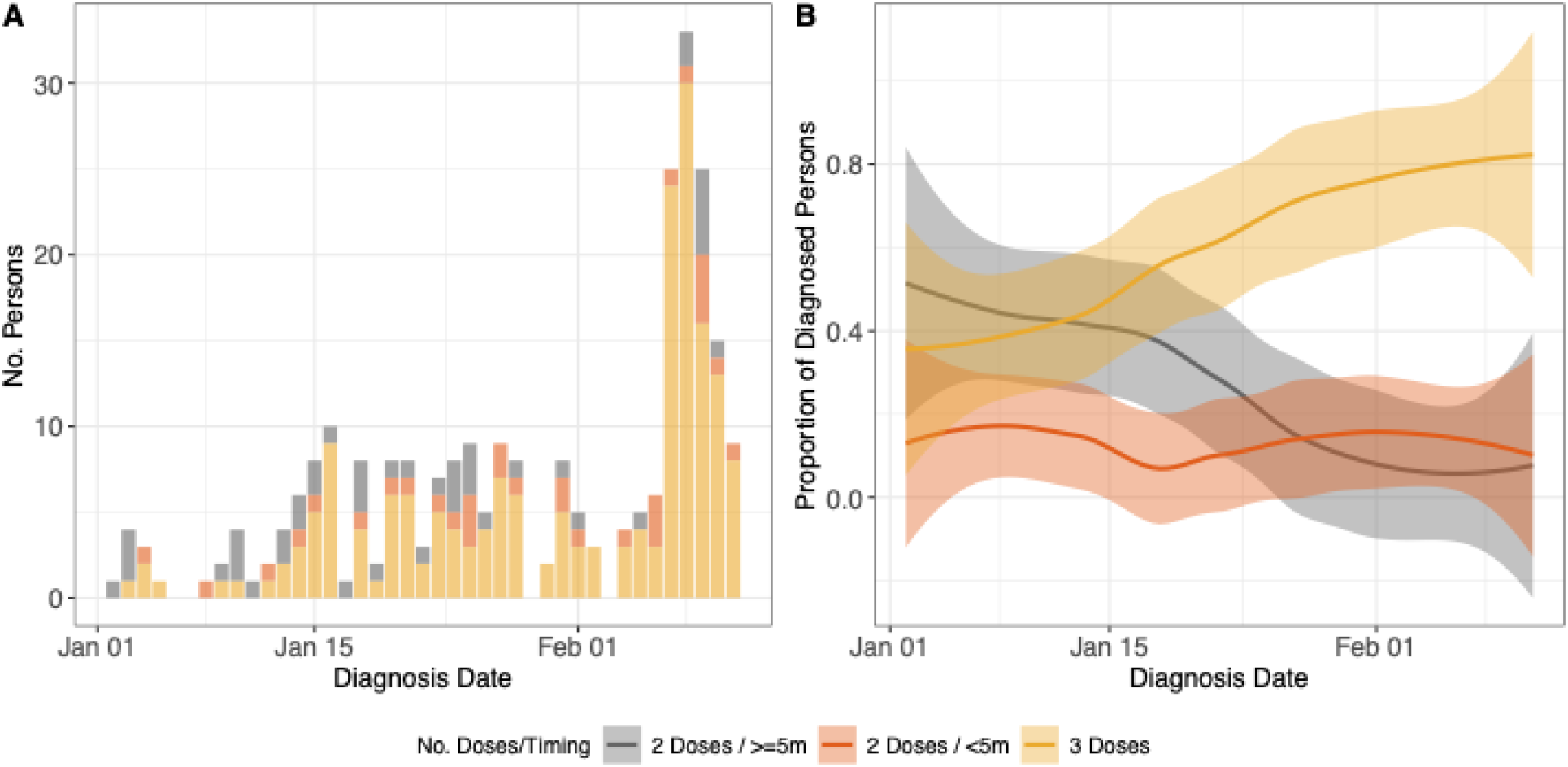
Number of and time since the last vaccine dose by diagnosis date, related to Table 2. N=263 persons included in the survival model analysis (Table 2). The diagnosis date is the date of the first positive or inconclusive test. **(A)** Number of persons in each number and timing of vaccine dose category over time by diagnosis date. **(B)** Smoothed proportion of total daily diagnosed persons in (A) belong to each number and timing of vaccine dose category over time.

